# LRRK2-Associated Parkinsonism With and Without *In Vivo* Evidence of Alpha-Synuclein Aggregates

**DOI:** 10.1101/2024.07.22.24310806

**Authors:** Lana M Chahine, David-Erick Lafontant, Seung Ho Choi, Hirotaka Iwaki, Cornelis Blauwendraat, Andrew B Singleton, Michael C Brumm, Roy N. Alcalay, Kalpana Merchant, Kelly Nicole Holohan Nudelman, Alain Dagher, Andrew Vo, Qin Tao, Charles S Venuto, Karl Kieburtz, Kathleen L Poston, Susan Bressman, Paulina Gonzalez-Latapi, Brian Avants, Christopher Coffey, Danna Jennings, Eduard Tolosa, Andrew Siderowf, Ken Marek, Tanya Simuni, Parkinson’s Progression Markers Initiative

## Abstract

**Background:** Among LRRK2-associated parkinsonism cases with nigral degeneration, over two-thirds demonstrate evidence of pathologic alpha-synuclein, but many do not. Understanding the clinical phenotype and underlying biology in such individuals is critical for therapeutic development. Our objective was to compare clinical and biomarker features, and rate of progression over 4 years follow-up, among LRRK2-associated parkinsonism cases with and without *in vivo* evidence of alpha-synuclein aggregates.

**Methods:** Data were from the Parkinson’s Progression Markers Initiative, a multicenter prospective cohort study. The sample included individuals diagnosed with Parkinson disease with pathogenic variants in LRRK2. Presence of CSF alpha-synuclein aggregation was assessed with seed amplification assay. A range of clinician– and patient-reported outcome assessments were administered. Biomarkers included dopamine transporter SPECT scan, CSF amyloid-beta_1-42_, total tau, phospho-tau_181_, urine bis(monoacylglycerol)phosphate levels, and serum neurofilament light chain. Linear mixed effects models examined differences in trajectory in CSF negative and positive groups.

**Results:** 148 LRRK2-parkinsonism cases (86% with G2019S variant), 46 negative and 102 positive for CSF alpha-synuclein seed amplification assay were included. At baseline, the negative group were older than the positive group (median [interquartile range] 69.1 [65.2-72.3] vs 61.5 [55.6-66.9] years, p<0.001) and a greater proportion were female (28 (61%) vs 43 (42%), p=0.035). Despite being older, the negative group had similar duration since diagnosis, and similar motor rating scale (16 [11-23] vs 16 [10-22], p=0.480) though lower levodopa equivalents. Only 13 (29%) of the negative group were hyposmic, compared to 75 (77%) of the positive group. Lowest putamen dopamine transporter binding expected for age and sex was greater in the negative vs positive groups (0.36 [0.29-0.45] vs 0.26 [0.22-0.37], p<0.001). Serum neurofilament light chain was higher in the negative group compared to the positive group (17.10 [13.60-22.10] vs 10.50 [8.43-14.70]; age-adjusted p-value=0.013). In terms of longitudinal change, the negative group remained stable in functional rating scale score in contrast to the positive group who had a significant increase (worsening) of 0.729 per year (p=0.037), but no other differences in trajectory were found.

**Conclusion:** Among individuals diagnosed with Parkinson disease with pathogenic variants in the LRRK2 gene, we found clinical and biomarker differences in cases without versus with *in vivo* evidence of CSF alpha-synuclein aggregates. LRRK2 parkinsonism cases without evidence of alpha-synuclein aggregates as a group exhibit less severe motor manifestations and decline may have more significant cognitive dysfunction. The underlying biology in LRRK2-parkinsonism cases without evidence of alpha-synuclein aggregates requires further investigation.

## Introduction

Individuals with LRRK2-associated parkinsonism uniformly demonstrate neuronal degeneration in the substantia nigra and locus coeruleus^1–3^, but the underlying proteinopathy is variable. A majority (60-80%) of cases demonstrate evidence of neuronal-predominant misfolded and aggregated alpha-synuclein (asyn), whether in vivo based on cerebrospinal fluid (CSF) testing or on post-mortem neuropathological examination^1,4,5^. However, over one-third may not have evidence of asyn aggregates. Understanding the clinical phenotype and underlying biology in such individuals is critical for molecularly-targeted therapeutic development^6^. Other pathologies present in some individuals with LRRK2-associated parkinsonism who do not demonstrate evidence of asyn aggregates include tauopathy, with Alzheimer’s disease (AD) type tau (3R and 4R) predominating, but some demonstrate hyperphosphorylated tau resembling progressive supranuclear palsy (PSP), and less commonly TAR DNA-binding protein 43 (TDP43)^3,7,8^.

Studies to date indicate that individuals with LRRK2-associated parkinsonism with and without evidence of asyn aggregates are largely clinically indistinguishable, with a few noted differences. Asyn positive LRRK2-parkinsonism cases have been reported to have more non-motor symptoms including hyposmia^4^, cognitive impairment, anxiety and orthostatic hypotension compared to asyn negative cases^2^. However, prior data are limited by small sample sizes and a lack of extensive clinical and biomarker characterization of cases. The Parkinson’s Progression Markers Initiative (PPMI) offers the unique opportunity to address key gaps in knowledge regarding clinical, biomarker, and genetic differences in LRRK2-associated parkinsonism with and without evidence of asyn aggregates, given that the cohort has had *in vivo* assessment of asyn aggregates in CSF as well as extensive longitudinal phenotyping in a relatively large number of cases. Indeed, findings have emerged from PPMI^4^ demonstrating that among individuals with LRRK2-associated parkinsonism, absence of detectable asyn aggregates is most prevalent among those who are normosmic, especially among females.

We undertook this study with the objectives of comparing among LRRK2-associated parkinsonism cases with and without evidence of asyn aggregates whether there are (1) differences in clinical features cross-sectionally and longitudinally (2) distinguishing features in available biofluid or imaging markers cross-sectionally and longitudinally (3) differences in prevalence of PD genetic risk. While acknowledging that LRRK2-associated parkinsonism without evidence of asyn aggregates are a biologically heterogeneous group, we hypothesized that LRRK2-associated parkinsonism without evidence of asyn aggregates would generally follow a more benign motor course.

## Materials and methods

### Sample

Data were from the PPMI, a multicenter prospective cohort study. PPMI methods have been described elsewhere in detail^9^. Briefly, PPMI recruited individuals diagnosed with PD based on clinical features who were sporadic (without known pathogenic variants associated with PD) and a group with parkinsonism and known pathogenic variants in LRRK2. Inclusion criteria for the sporadic PD group were abnormal dopamine transporter (DAT) SPECT imaging by visual inspection, 2 years or less since diagnosis, not receiving dopaminergic treatment and not expected to require it within 6 months of enrollment. The LRRK2-associated parkinsonism group was enrolled irrespective of treatment and if disease duration was 7 or less years. Exclusion criteria for all enrolled groups included dementia and medical conditions that preclude study activities.

The sample for this analysis is comprised of individuals with LRRK2-associated parkinsonism (LRRK2-parkinsonism) and a sporadic PD (sPD) group frequency matched to the LRRK2-parkinsonism group for age and time since diagnosis at enrollment.

Inclusion criteria for this analysis were: (1) availability of asyn seed amplification assay (SAA) result (see methods below) (2) positive asyn SAA (CSFasynSAA+) result for the matched sPD group. Exclusion criteria were lowest putamen DAT specific binding ratio ≥65% of expected for age and sex in individuals who had a negative asyn SAA (CSFasynSAA-) result, presence of known pathogenic *GBA1* variant (as presence of pathogenic glucocerebrosidase (*GBA1*) variants in individuals with LRRK2 can potentially modify the phenotype), and inconclusive or multiple system atrophy-like SAA results.

Baseline visit (time zero) for this analysis was the baseline study assessment for participants in the LRRK2 parkinsonism group and for the sPD group it was the first visit at which they were frequency matched for age and time since diagnosis.

### Assessments of Motor and Non-Motor Function

Motor and non-motor assessment of signs, symptoms and function in PPMI that are assessed at baseline and at each annual visit are as follows:

– Demographics: age, sex at birth, years of education, self-reported race and ethnicity
– Clinical history: age at parkinsonism symptom onset, duration since PD clinical diagnosis at baseline visit, levodopa equivalent daily dose (LEDD)
– Movement Disorders Society Modified Unified Parkinson’s Disease Rating Scale (MDS-UPDRS) parts 1, 2, and 3. An ambulatory capacity score was calculated as the sum of MDS-UPDRS items 2.12, 2.13, 3.10, 3.11, 3.12. Medication OFF part 3 scores were missing on a substantial portion of participants and only medication ON state scores are included in this analysis
– Modified Schwab and England
– Cognitive assessment: the Montreal Cognitive Assessment (MoCA) and the following neuropsychological tests were administered to assess the respective specified domains: Hopkins Verbal Learning Test—Revised (HVLT-R)^10^; visuospatial function: Benton Judgment of Line Orientation 15-item (split-half) version^11^, and executive function along with working memory: Letter-Number Sequencing and semantic (animal) fluency^12^. Published norms were applied, as referenced.
– Psychiatric assessments: Geriatric Depression Scale-15 item (GDS-15), State and Trait anxiety scale (STAI), Epworth Sleepiness Scale (ESS), Questionnaire for Impulsive-Compulsive Disorders in Parkinson’s Disease–Rating Scale (QUIP-RS)
– Other non-motor: REM sleep behavior disorder questionnaire; possible RBD defined as RBDSQ≥6, Scales for Outcomes in Parkinson’s-Autonomic (SCOPA-AUT)
– Olfactory function is assessed with the 38 item University of Pennsylvania Smell Identification Test (UPSIT). Hyposmia is defined as UPSIT score in the ≤15 percentile expected for age and sex^13^.

### Genotyping

Genotyping methods in PPMI are described in detail at ppmi-info.org. Briefly, each PPMI participant receives a determination of presence or absence of pathogenic variants in the LRRK2 gene (or other genes) as well as APOE genotype. Population genetic structure was inferred with principal component analysis as described^14^.

In addition, we procured genome sequencing data from the Accelerating Medicines Partnership Parkinson’s Disease (AMP-PD) project. The data processing methodology is detailed in a public GitHub repository^15^, follows the methods outlined by Nalls et al^16^, utilizing 90 risk-associated SNPs. However, for this study, we omitted two SNPs located in the LRRK2 region. We thus generated a modified polygenic risk score (mPRS), the cumulative risk weighted by the effect estimates of associated genetic variants, consisting of 88 SNPs.

### Biomarker assessments

Presence of aggregated alpha-synuclein in CSF obtained at the baseline visit was assessed using the alpha-synuclein (asyn) seed amplification assay (SAA) as described^4,17^. The Fmax (highest raw fluorescence from each well), T50 (time to reach 50% of the Fmax), and TTT (time to reach a target RFU threshold) were used to define positive (CSFasynSAA+), inconclusive, negative (CSFasynSAA-), and multiple system atrophy-like (MSA-like) assays as described^4,17^.

Dopamine transporter binding (DAT) was assessed with DATscan and SPECT as previously described^8^. Percent of expected lowest putamen specific binding ratio (SBR) for age and sex was determined using normative data from healthy controls in PPMI.

Other available biomarkers measured in CSF or serum were amyloid-beta_1-42_, total tau, phospho-tau_181_, and serum neurofilament light (NfL) chain, with immunoassays, as described^18,19^. Amyloid-beta_1-42_, total tau, phospho-tau_181_, levels were categorized as abnormal based on conventional Alzheimer’s disease (AD) cutoffs^20^ as follows: CSF amyloid-beta_1-42_ ≤ 683, total tau ≥ 266, and CSF phospho-tau_181_ ≥ 24. In addition, we examined cutoffs modified for the PD population^21^ as follows: CSF amyloid-beta_1-42_ ≤ 710, total tau ≥ 148, and CSF phospho-tau_181_ ≥ 13.

Assessment of urine bis(monoacylglycero)phosphate (BMP) isforms (total di-18:1 BMP, total di-22:6-BMP, and 2,2’ di-22:6 BMP) was performed by Nextcea, Inc. (Woburn, MA) using targeted ultra-performance liquid chromatography-mass spectrometry as described^22^.

### Statistical Analysis

Baseline demographic and clinical features were compared in the CSFasynSAA– and CSFasynSAA+ using two-sample Wilcoxon rank sum test, chi-square test or Fisher’s exact test as appropriate. To account for differences due to age, linear regression and logistic regression adjusting for age for continuous and categorical outcomes, respectively were used to model clinical outcomes and biomarkers with SAA as an explanatory variable. Log, square root, or rank transformations were applied to models with non-normally distributed residuals. The specific transformations used were marked on the tables and detailed in the table footers. Summary statistics were examined for motor, non-motor, and biologic variables from baseline to year 4.

Only individuals with at least 1 annual follow-up visit following baseline were included in longitudinal analyses. To assess whether the longitudinal trajectory of the outcome measures differed between CSFasynSAA– and CSFasynSAA+ groups, generalized linear mixed-effects models (LMM) with random intercept and slope and unstructured working correlation structure were employed. Specifically, CSF asyn SAA status, time in years, and their interaction were included in the models. This analysis assumed a linear fit in the link function of mean responses over time from year 1 to year 4, wherever available, using the Restricted Maximum Likelihood and Residual Pseudo-Likelihood methods when appropriate. Continuous biologic CSF outcomes were ranked at each time point and modeled to evaluate whether the longitudinal trajectory of the mean rank response differed by CSF asyn SAA groups, assuming a linear fit in the mean rank of each response over time from year 1 to year 4, when available. Similarly, models were employed to assess whether the longitudinal trajectory of log odds for categorical response variables differed based on CSF asyn SAA status from year 1 to year 4. Random intercept only models were used for outcomes with convergence issues. Wald tests were conducted to assess the statistical significance of the interaction term between CSF asyn SAA status and time. A quadratic fit model was also tested if the linear fit did not result in a significant interaction. To explore sex differences, a three-way interaction model with sex, CSF asyn SAA, and time was also tested. An identity link and logit link were chosen for continuous and categorical response variables, respectively. Time effect p-values were reported for all models, with separate time effects provided for each CSF asyn SAA status when the interaction term was significant.

All models adjusted for baseline value of the outcome, age, sex, years since diagnosis at enrollment, and genetic principal components PC1, PC2, PC3^14^. Models involving outcomes that may be affected by PD medications, such as MDS-UPDRS Part 3, and Ambulatory Capacity Score also adjusted for time-varying LEDD in the model.

All longitudinal analyses were conducted under the assumption of missing at random (MAR). Sensitivity analyses were employed to evaluate the plausibility of the MAR assumption. Intermittent missing values were imputed using Monte Carlo Markov Chain methods^23^. Multiple imputation was used for outcomes displaying significant interactions. Notably, a one-dimensional tipping point analysis was utilized to assess the significance of the interaction term by systematically shifting the mean of missing values at year 4 for these outcomes in the opposite direction of significance, identifying the point at which the interaction term becomes nonsignificant.

To determine if differences in CSFasynSAA– and CSFasynSAA+ parkinsonism cases vary according to LRRK2 status, when cross-sectional or longitudinal analysis revealed significant differences in the LRRK2 parkinsonism CSFasynSAA– and CSFasynSAA+ group for a given outcome, the outcome was then compared in the LRRK2 parkinsonism CSFasynSAA-group to the sPD CSFasynSAA+ group using the same statistical method.

For comparison of genetic risk variants in the LRRK2-associated parkinsonism cases, we compared mPRS in CSFasynSAA+ and CSFasynSAA-groups using logistic regression with CSFasynSAA-as the reference group. In addition, we conducted an examination of individual GWAS risk variants to evaluate their association with CSF asyn SAA status, adjusting for age, sex, and the first three genetic principal components. Given the exploratory nature of this study, we set the significance threshold at 0.05 (two-tailed).

All analyses were conducted in SAS Institute Inc. (SAS Institute Inc version 9.4 Cary, NC).

## Results

### Sample Characteristics

PPMI enrolled 184 individuals with LRRK2-associated parkinsonism. 36 were excluded (reasons for exclusion: no CSF asyn SAA result available (n=17), GBA1 pathogenic variant present (n=8), CSFasynSAA– and DAT-(n=9), CSF asyn SAA inconclusive or MSA-like (n=2)).

The final analytic sample included 148 LRRK2-associated parkinsonism cases and a comparator group of 378 sporadic PD CSFasynSAA+ (sPD) frequency matched to them by age and disease duration. Seven participants did not have follow-up beyond baseline. Up to 4 follow-up visits (after baseline) were expected for 141 LRRK2-associated parkinsonism cases; the majority completed year 4 (31 (69%) CSFasynSAA– and 78 (81%) CSFasynSAA+cases). Among the 32 cases who did not complete year 4, 9 contributed data at later time points, 13 withdrew from the study before year 4, and 7 were lost to follow-up.

Baseline characteristics of the sample are shown in Table 1. Among the LRRK2-associated parkinsonism cases, 46 (31%) were CSFasynSAA– and 102 (69%) were CSFasynSAA+. The LRRK2 CSFasynSAA-group, compared to the CSFasynSAA+ group, were older at first study visit (median [IQR] 69.1 [65.2-72.3] vs 61.5 [55.6-66.9] years, p<0.001), had older age of symptom onset (64.6 [58.5-68.8] vs 57.6 [49.0-62.1] years, p<0.001), and were more likely to be female (61% vs 42%, p=0.035), but they had similar duration since clinical diagnosis (1.9 [0.9-4.2] vs 2.3 [1.3-4.6] years, p=0.288). While the majority of pathogenic variants were G2019S (86%), among the 20 cases that were not LRRK2 G2019S, R1441G was the most common, and 12/17 (71%) were in the CSFasynSAA-group.

**Table 1:** Sample demographics and other characteristics.

| Variable |  |  |  | p-values |  |  |
| --- | --- | --- | --- | --- | --- | --- |
|  | 1. LRRK2 SAA- (N=46) | 2. LRRK2 SAA+ (N=102) | 3. sPD SAA+ (N=378) | 1 vs 2 | 1 vs 3 | 2 vs 3 |
| Age at baseline, years, Median [IQR] | 69.1 [65.2-72.3] | 61.5 [55.6-66.9] | 62.1 [57.9-65.6] | <0.001 | <0.001 | 0.710 |
| Age at PD onset, years, Median [IQR] | 64.6 [58.5-68.8] | 57.6 [49.0-62.1] | 57.9 [53.0-61.3] | <0.001 | <0.001 | 0.591 |
| Male sex, N (%) | 18 (39%) | 59 (58%) | 242 (64%) | 0.035 | 0.001 | 0.252 |
| Years of education, Median [IQR] | 14.5 [10.0-17.0] | 17.0 [14.0-19.0] | 16.0 [14.0-18.0] | 0.001 | 0.003 | 0.067 |
| Years since PD diagnosis, Median [IQR] | 1.9 [0.9-4.2] | 2.3 [1.3-4.6] | 2.6 [0.8-5.3] | 0.288 | 0.465 | 0.782 |
| Race (% White), N (%) | 40 (87%) | 96 (94%) | 352 (94%) | 0.192 | 0.113 | 0.925 |
| Hispanic, N (%) | 12 (26%) | 15 (15%) | 6 (2%) | 0.097 | <0.001 | <0.001 |
| LED, Median [IQR] | 205 [100-385] | 500 [300-765] | 300 [0-580] | <0.001 | 0.459 | <0.001 |
| LED=0, N (%) | 7 (16%) | 7 (7%) | 122 (36%) | 0.125 | 0.009 | <0.001 |
| LRRK2 Variant#, N (%) |  |  |  | <0.001 |  |  |
| G2019S | 33 (72%) | 95 (93%) | N/A |  |  |  |
| N1437H | 0 (0%) | 1 (1%) | N/A |  |  |  |
| R1441G | 12 (26%) | 5 (5%) | N/A |  |  |  |
| R1441C | 0 (0%) | 1 (1%) | N/A |  |  |  |
| I2020T | 1 (2%) | 0 (0%) | N/A |  |  |  |
| APOE Genotype - number of e4 alleles#, N (%) |  |  |  | 0.307 | 0.974 | 0.142 |
| 0 e4 alleles | 32 (73%) | 78 (80%) | 137 (72%) |  |  |  |
| 1 e4 allele | 11 (25%) | 18 (19%) | 48 (25%) |  |  |  |
| 2 e4 alleles | 1 (2%) | 1 (1%) | 4 (2%) |  |  |  |

LED=levodopa equivalent daily dose (mg)
Missing data in LRRK2 parkinsonism cases: Age at PD onset, n=10 (6.8%); Years since PD diagnosis, n=4 (2.7%); LED, n=3 (2.0%); APOE Genotype, n=7 (4.7%).

### Baseline Motor and Non-Motor Features

Table 2 shows baseline motor and non-motor measures. Despite being older, having similar duration since clinical diagnosis at baseline assessment, and having significantly lower LEDD (median [IQR] 205 [100-385] vs 500 [300-765], p<0.001), the LRRK2 CSFasynSAA-group had similar scores to the LRRK2 CSFasynSAA+ group in MDS-UPDRS total score and subscores, including part III ON score (median [IQR] 16 [11-23] vs 16 [10-22], p=0.480).

**Table 2:**
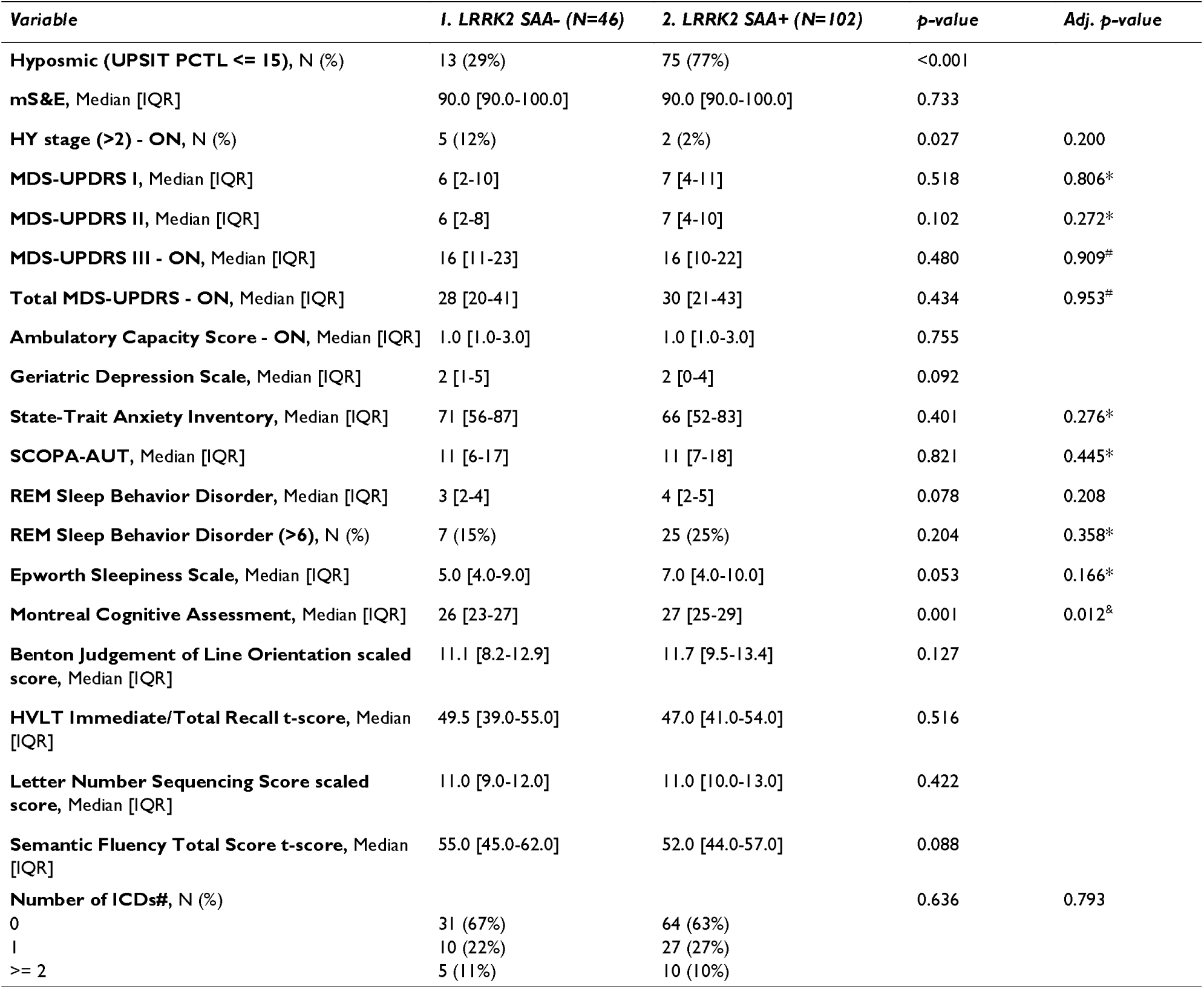
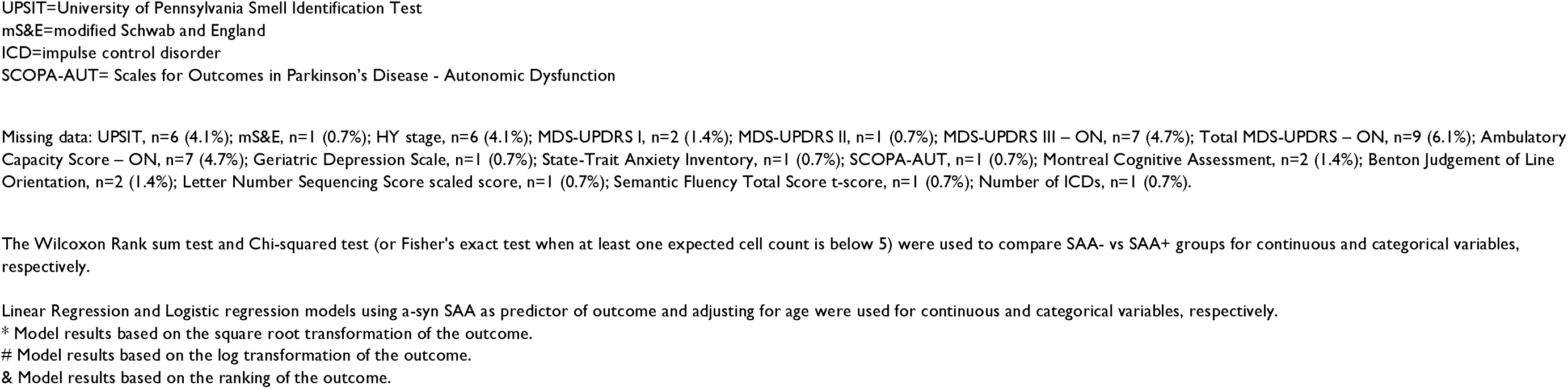
Comparison of motor and non-motor features in LRRK2 parkinsonism CSF asyn SAA– and SAA+ cases.

Only 13 (29%) of the LRRK2 CSFasynSAA-group were hyposmic, compared to 75 (77%) of the LRRK2 CSFasynSAA+ group.

MoCA total score was lower in the LRRK2 CSFasynSAA-group compared to the LRRK2 aSyn-CSFasynSAA+ (median [IQR] 26 [23-27] vs 27 [25-29], p=0.001), but this did not remain significant after adjusting for age, sex, and education (p=0.064). MoCA score was also lower in the LRRK2 CSFasynSAA-group compared to the sPD group (median [IQR] 26 [23-27] vs 28 [26-29], unadjusted p<0.001; adjusted for age, sex, and education p-value= 0.005).

There were no differences in other non-motor measures or tests of cognitive function in the two groups (Table 2).

### Baseline Imaging and Biofluid Biomarker Assessments

Median [IQR] lowest putamen DAT SBR expected for age and sex in the LRRK2 CSFasynSAA-group (0.36 [0.29-0.45]) was significantly greater than in the LRRK2 CSFasynSAA+ (0.26 [0.22-0.37], p<0.001; Table 3) but not the sPD group (0.34 [0.25-0.42]; p=0.101).

**Table 3:** Comparison of imaging and biofluid biomarkers at baseline in LRRK2 parkinsonism CSF asyn SAA– and SAA+ cases.

| Variable | LRRK2 SAA- (N=46) | LRRK2 SAA+ (N=102) | p-value | Adj. p-value |
| --- | --- | --- | --- | --- |
| **DAT SBR lowest putamen**, Median [IQR] | 0.36 [0.29-0.45] | 0.26 [0.22-0.37] | <.001 |  |
| **CSF abeta<sup>†</sup>**, Median [IQR] | 928.3 [657.9-1,181.5] | 793.4 [605.7-1,050.7] | 0.160 | 0.181<sup>&</sup> |
| **CSF abeta ≤ 683**, N (%) | 12 (29%) | 32 (33%) | 0.607 | 0.632 |
| **CSF abeta ≤ 710**, N (%) | 12 (29%) | 38 (39%) | 0.232 | 0.238 |
| **CSF total tau<sup>†</sup>**, Median [IQR] | 186.3 [135.5-229.3] | 148.4 [118.6-193.6] | 0.004 | 0.136<sup>&</sup> |
| **CSF tau ≥ 266**, N (%) | 6 (14%) | 7 (7%) | 0.216 | 0.862 |
| **CSF tau ≥ 148**, N (%) | 32 (74%) | 50 (51%) | 0.010 | 0.144 |
| **CSF ptau<sup>†</sup>**, Median [IQR] | 15.1 [11.5-18.6] | 12.7 [10.0-15.6] | 0.003 | 0.160<sup>&</sup> |
| **CSF ptau ≥ 24**, N (%) | 4 (9%) | 6 (6%) | 0.493 | 0.988 |
| **CSF ptau ≥ 13**, N (%) | 30 (70%) | 47 (48%) | 0.017 | 0.304 |
| **CSF tau-abeta Ratio<sup>†</sup>**, Median [IQR] | 0.193 [0.163-0.231] | 0.177 [0.159-0.204] | 0.186 | 0.931<sup>&</sup> |
| **Serum NfL**, Median [IQR] | 17.10 [13.60-22.10] | 10.50 [8.43-14.70] | <.001 | 0.013 |
| **Total di-18:1 BMP**, Median [IQR] | 15 [7-29] | 11 [7-21] | 0.204 | 0.114<sup>#</sup> |
| **Total di-22:6-BMP**, Median [IQR] | 77 [45-108] | 59 [39-97] | 0.196 | 0.337<sup>#</sup> |
| **2.2 di-22:6 BMP**, Median [IQR] | 60 [38-90] | 47 [28-80] | 0.216 | 0.472<sup>#</sup> |

DAT SBR lowest putamen=dopamine transporter specific binding ratio, percent expected for age and sex, lowest of the right or left putamen values
Missing data: DAT SBR lowest putamen, n=8 (5.4%); CSF abeta, n=9 (6.1%); CSF tau and ptau, n=7 (4.7%); Serum NfL: n=27 (18.2%); Urine BMP, n=14 (9.5%).
<sup>†</sup>Scores were imputed with their upper and lower limits of detection.
The Wilcoxon Rank sum test and Chi-squared test (or Fisher's exact test when at least one expected cell count is below 5) were used to compare SAA- vs SAA+ groups for continuous and categorical variables, respect Linear Regression and Logistic regression models using a-syn SAA as predictor of outcome and adjusting for age were used for continuous and categorical variables, respectively.
<sup>#</sup>Model results based on the log transformation of the outcome.
<sup>&</sup> Model results based on the ranking of the outcome.

Median [IQR] serum NfL was significantly higher in the LRRK2 CSFasynSAA-(17.10 [13.60-22.10]) compared to the LRRK2 CSFasynSAA+ group (10.50 [8.43-14.70], p<0.001) and the sPD group (12.60 [9.60-16.10], p<0.001). Differences in LRRK2 CSFasynSAA– and CSFasynSAA+ serum NfL remained significant after adjusting for age (p=0.013). CSF total tau and phospho-tau tended to be higher in the LRRK2 CSFasynSAA-group compared to the LRRK2 CSFasynSAA+ group, but the results did not remain significant once adjusting for age (Table 3). Otherwise, no biofluid biomarkers differed between the groups.

### Comparison of PD genetic risk variants

The analysis comparing risk variants was confined to LRRK2-parkinsonism cases of European ancestry (n=130) of which 48 were CSFasynSAA– and 82 were CSFasynSAA+. The analysis did not reveal a statistically significant association between mPRS and SAA status (Odds Ratio: 0.78 [95% Wald Confidence Interval: 0.52, 1.19], p=0.25). However, in the individual variant analysis, three risk-associated variants emerged as noteworthy: rs11557080 (p=0.0034), located in the 3’ UTR of RAB29; rs12951632 (p=0.026), an intron variant of RETREG3; and rs6808178 (p=0.038), an intron variant of LINC00693.

### Longitudinal Change in Motor and Non-Motor Features and Biomarkers

Raw mean values at each follow-up time point in the CSFasynSAA+ and CSFasynSAA-groups are shown in Table 4. Results of the LMMs are shown in supplementary table 1. The MDS-UPDRS II score did not significantly change over time in the CSFasynSAA-group (β=0.108 (95% Wald CI: –0.466, 0.682, p=0.711) whereas in the CSFasyn SAA+ group, it increased significantly by 0.837 points per year (95% Wald CI: 0.467, 1.207, p<0.001). Thus, despite the CSFasynSAA-group being older at enrollment and having lower LED, the CSFasynSAA+ group worsened by 0.729 points more per year compared to the CSFasyn SAA-group (p=0.037). Tipping-point analysis showed that imputed MDS-UPDRS II for participants in the CSFasynSAA+ group who had missing data up until (including) year 4 would have to be approximately 2-points lower on average in the CSFasynSAA+ group to nullify the significance of the main effect.

**Table 4:**
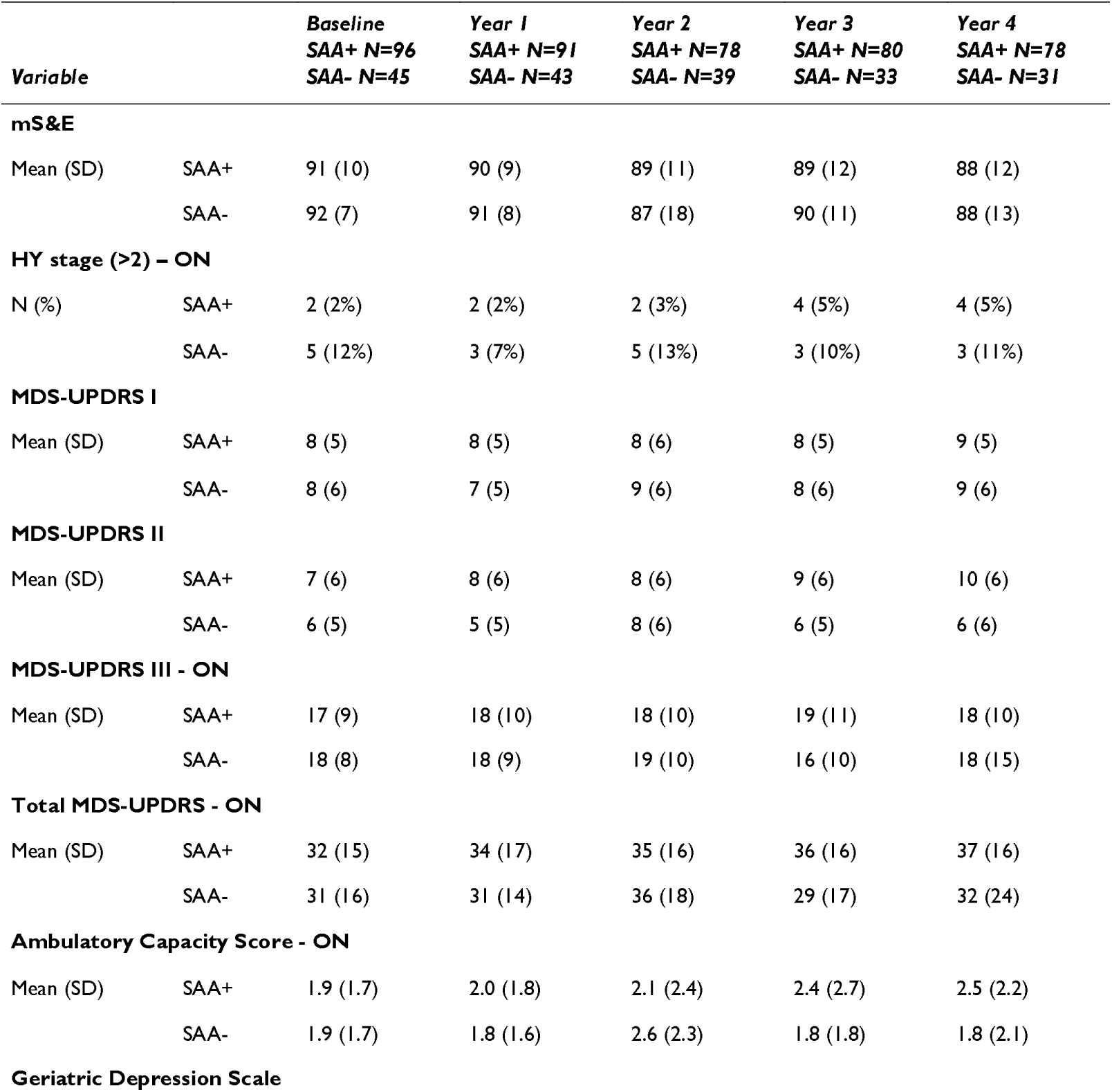
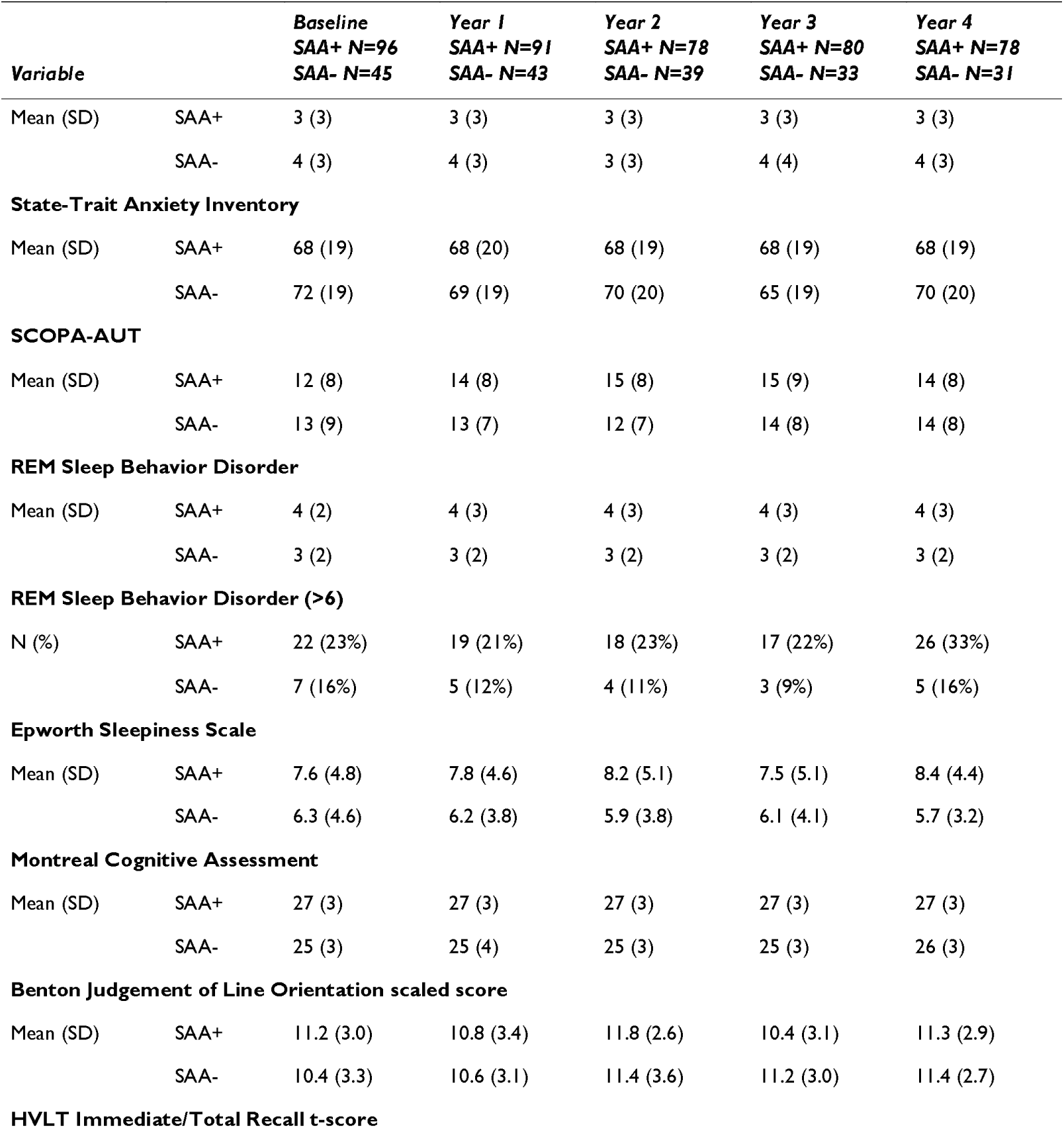
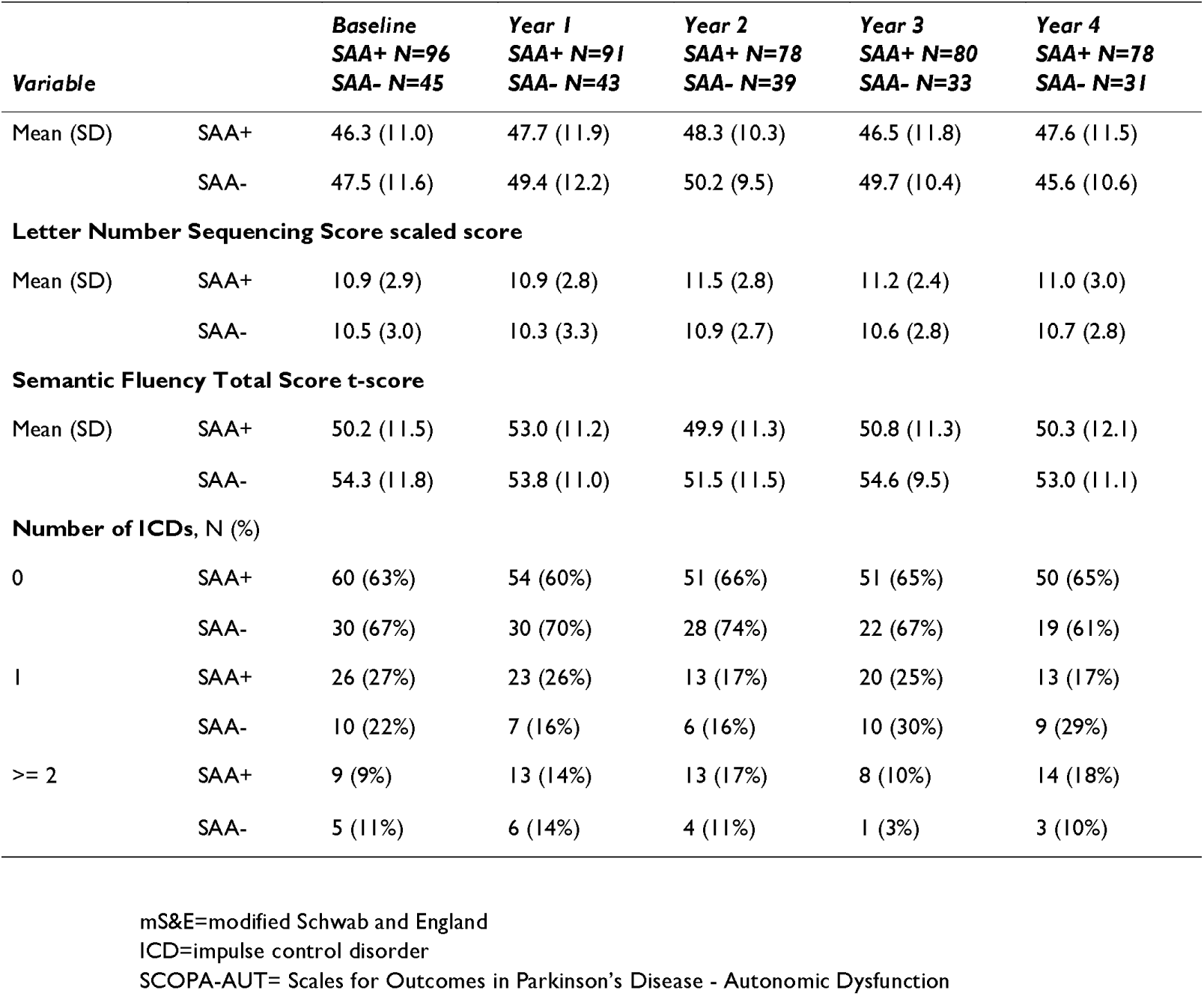
Longitudinal assessment of motor and non-motor features.

|  |  | Baseline<br>SAA+ N=96<br>SAA- N=45 | Year 1<br>SAA+ N=91<br>SAA- N=43 | Year 2<br>SAA+ N=78<br>SAA- N=39 | Year 3<br>SAA+ N=80<br>SAA- N=33 | Year 4<br>SAA+ N=78<br>SAA- N=31 |
| --- | --- | --- | --- | --- | --- | --- |
| Variable |  |  |  |  |  |  |
| mS&E |  |  |  |  |  |  |
| Mean (SD) | SAA+ | 91 (10) | 90 (9) | 89 (11) | 89 (12) | 88 (12) |
|  | SAA- | 92 (7) | 91 (8) | 87 (18) | 90 (11) | 88 (13) |
| HY stage (>2) – ON |  |  |  |  |  |  |
| N (%) | SAA+ | 2 (2%) | 2 (2%) | 2 (3%) | 4 (5%) | 4 (5%) |
|  | SAA- | 5 (12%) | 3 (7%) | 5 (13%) | 3 (10%) | 3 (11%) |
| MDS-UPDRS I |  |  |  |  |  |  |
| Mean (SD) | SAA+ | 8 (5) | 8 (5) | 8 (6) | 8 (5) | 9 (5) |
|  | SAA- | 8 (6) | 7 (5) | 9 (6) | 8 (6) | 9 (6) |
| MDS-UPDRS II |  |  |  |  |  |  |
| Mean (SD) | SAA+ | 7 (6) | 8 (6) | 8 (6) | 9 (6) | 10 (6) |
|  | SAA- | 6 (5) | 5 (5) | 8 (6) | 6 (5) | 6 (6) |
| MDS-UPDRS III - ON |  |  |  |  |  |  |
| Mean (SD) | SAA+ | 17 (9) | 18 (10) | 18 (10) | 19 (11) | 18 (10) |
|  | SAA- | 18 (8) | 18 (9) | 19 (10) | 16 (10) | 18 (15) |
| Total MDS-UPDRS - ON |  |  |  |  |  |  |
| Mean (SD) | SAA+ | 32 (15) | 34 (17) | 35 (16) | 36 (16) | 37 (16) |
|  | SAA- | 31 (16) | 31 (14) | 36 (18) | 29 (17) | 32 (24) |
| Ambulatory Capacity Score - ON |  |  |  |  |  |  |
| Mean (SD) | SAA+ | 1.9 (1.7) | 2.0 (1.8) | 2.1 (2.4) | 2.4 (2.7) | 2.5 (2.2) |
|  | SAA- | 1.9 (1.7) | 1.8 (1.6) | 2.6 (2.3) | 1.8 (1.8) | 1.8 (2.1) |
| Geriatric Depression Scale |  |  |  |  |  |  |

|  |  | Baseline | Year 1 | Year 2 | Year 3 | Year 4 |
| --- | --- | --- | --- | --- | --- | --- |
|  |  | SAA+ N=96 | SAA+ N=91 | SAA+ N=78 | SAA+ N=80 | SAA+ N=78 |
| Variable |  | SAA- N=45 | SAA- N=43 | SAA- N=39 | SAA- N=33 | SAA- N=31 |
| Mean (SD) | SAA+ | 3 (3) | 3 (3) | 3 (3) | 3 (3) | 3 (3) |
|  | SAA- | 4 (3) | 4 (3) | 3 (3) | 4 (4) | 4 (3) |
| State-Trait Anxiety Inventory |  |  |  |  |  |  |
| Mean (SD) | SAA+ | 68 (19) | 68 (20) | 68 (19) | 68 (19) | 68 (19) |
|  | SAA- | 72 (19) | 69 (19) | 70 (20) | 65 (19) | 70 (20) |
| SCOPA-AUT |  |  |  |  |  |  |
| Mean (SD) | SAA+ | 12 (8) | 14 (8) | 15 (8) | 15 (9) | 14 (8) |
|  | SAA- | 13 (9) | 13 (7) | 12 (7) | 14 (8) | 14 (8) |
| REM Sleep Behavior Disorder |  |  |  |  |  |  |
| Mean (SD) | SAA+ | 4 (2) | 4 (3) | 4 (3) | 4 (3) | 4 (3) |
|  | SAA- | 3 (2) | 3 (2) | 3 (2) | 3 (2) | 3 (2) |
| REM Sleep Behavior Disorder (>6) |  |  |  |  |  |  |
| N (%) | SAA+ | 22 (23%) | 19 (21%) | 18 (23%) | 17 (22%) | 26 (33%) |
|  | SAA- | 7 (16%) | 5 (12%) | 4 (11%) | 3 (9%) | 5 (16%) |
| Epworth Sleepiness Scale |  |  |  |  |  |  |
| Mean (SD) | SAA+ | 7.6 (4.8) | 7.8 (4.6) | 8.2 (5.1) | 7.5 (5.1) | 8.4 (4.4) |
|  | SAA- | 6.3 (4.6) | 6.2 (3.8) | 5.9 (3.8) | 6.1 (4.1) | 5.7 (3.2) |
| Montreal Cognitive Assessment |  |  |  |  |  |  |
| Mean (SD) | SAA+ | 27 (3) | 27 (3) | 27 (3) | 27 (3) | 27 (3) |
|  | SAA- | 25 (3) | 25 (4) | 25 (3) | 25 (3) | 26 (3) |
| Benton Judgement of Line Orientation scaled score |  |  |  |  |  |  |
| Mean (SD) | SAA+ | 11.2 (3.0) | 10.8 (3.4) | 11.8 (2.6) | 10.4 (3.1) | 11.3 (2.9) |
|  | SAA- | 10.4 (3.3) | 10.6 (3.1) | 11.4 (3.6) | 11.2 (3.0) | 11.4 (2.7) |
| HVLT Immediate/Total Recall t-score |  |  |  |  |  |  |

| **Variable** |  | **Baseline**<br>**SAA+ N=96**<br>**SAA- N=45** | **Year 1**<br>**SAA+ N=91**<br>**SAA- N=43** | **Year 2**<br>**SAA+ N=78**<br>**SAA- N=39** | **Year 3**<br>**SAA+ N=80**<br>**SAA- N=33** | **Year 4**<br>**SAA+ N=78**<br>**SAA- N=31** |
| --- | --- | --- | --- | --- | --- | --- |
| Mean (SD) | SAA+ | 46.3 (11.0) | 47.7 (11.9) | 48.3 (10.3) | 46.5 (11.8) | 47.6 (11.5) |
|  | SAA- | 47.5 (11.6) | 49.4 (12.2) | 50.2 (9.5) | 49.7 (10.4) | 45.6 (10.6) |
| **Letter Number Sequencing Score scaled score** |  |  |  |  |  |  |
| Mean (SD) | SAA+ | 10.9 (2.9) | 10.9 (2.8) | 11.5 (2.8) | 11.2 (2.4) | 11.0 (3.0) |
|  | SAA- | 10.5 (3.0) | 10.3 (3.3) | 10.9 (2.7) | 10.6 (2.8) | 10.7 (2.8) |
| **Semantic Fluency Total Score t-score** |  |  |  |  |  |  |
| Mean (SD) | SAA+ | 50.2 (11.5) | 53.0 (11.2) | 49.9 (11.3) | 50.8 (11.3) | 50.3 (12.1) |
|  | SAA- | 54.3 (11.8) | 53.8 (11.0) | 51.5 (11.5) | 54.6 (9.5) | 53.0 (11.1) |
| **Number of ICDs, N (%)** |  |  |  |  |  |  |
| 0 | SAA+ | 60 (63%) | 54 (60%) | 51 (66%) | 51 (65%) | 50 (65%) |
|  | SAA- | 30 (67%) | 30 (70%) | 28 (74%) | 22 (67%) | 19 (61%) |
| 1 | SAA+ | 26 (27%) | 23 (26%) | 13 (17%) | 20 (25%) | 13 (17%) |
|  | SAA- | 10 (22%) | 7 (16%) | 6 (16%) | 10 (30%) | 9 (29%) |
| >= 2 | SAA+ | 9 (9%) | 13 (14%) | 13 (17%) | 8 (10%) | 14 (18%) |
|  | SAA- | 5 (11%) | 6 (14%) | 4 (11%) | 1 (3%) | 3 (10%) |

mS&E=modified Schwab and England
ICD=impulse control disorder
SCOPA-AUT= Scales for Outcomes in Parkinson's Disease - Autonomic Dysfunction

None of the other assessed rating scales, imaging, or biofluid biomarkers changed significantly in the two groups when the outcome was modeled as linear. When a quadratic term was introduced, the interaction with the second order term was significant for SCOPA-AUT, though there was minimal overall change in SCOPA-AUT total score (Table 4).

None of the imaging, CSF, serum, or urine biomarkers changed differently in the CSFasynSAA– and CSFasynSAA+ groups over time (supplementary table 2). Data on biofluid biomarkers were missing on a substantial number at later time points of follow-up (supplementary table 2).

Testing of a 3-way interaction term between sex and SAA status did not reveal any differences in change according to SAA and sex (supplementary table 1), but sample sizes in the subgroups at later time points were small.

## Discussion

In this large sample of individuals with LRRK2-associated parkinsonism, we compared clinical, imaging and biofluid biomarker, and genetic characteristics among those with evidence of CSF asyn aggregates compared to those without. Importantly, and unique to this cohort, all assessments occurred *in vivo* in participants who had received a clinical diagnosis of PD and had dopaminergic dysfunction as evidenced by DAT imaging. Taken together, our results indicate that while the CSF asyn CSFasynSAA– and CSFasynSAA+ groups are largely similar, there are some important differences. The CSF asyn CSFasynSAA-group had less severe motor dysfunction (and a trend toward more severe cognitive dysfunction at baseline). Concordantly, they had less advanced dopaminergic neuron dysfunction, as evidenced by DAT binding measures. By contrast, the CSFasynSAA-group had higher serum NfL, a biomarker that predicts increased risk of cognitive decline^24^. Interpretation of these results requires consideration for sex and age differences in the compared groups, as well as differences in disease duration at enrollment. Longitudinal analysis revealed that the CSFasynSAA-group, despite being older and receiving less dopaminergic therapy, did not decline in motor functional rating scale, in contrast to the CSFasynSAA+ group who had significant worsening of functional impairment over time.

### High prevalence of LRRK2 parkinsonism cases without evidence for asyn aggregates

In the PPMI sample of LRRK2 parkinsonism cases included in this analysis, one-third had no evidence of asyn aggregates based on CSF asyn SAA. This is in contrast to sporadic PD— individuals with a clinical diagnosis of PD who do not have any known pathogenic variants— where only 6.7%^4^.-9%^4,25^ of cases do not have evidence of asyn aggregates.

It is likely that most cases that are negative for CSF asyn SAA are negative for asyn in the brain. This is supported by several lines of evidence including measurement of asyn with a variety of methods and autopsy-CSF correlation^26^, including one of the cases included in this analysis that was examined postmortem and showed no Lewy pathology^4^. Nevertheless, it is possible that in some cases the CSF test is false negative. Indeed, some neuropathologically examined cases with confirmed Lewy body pathology have been CSF asyn SAA-; these are most often focal Lewy pathology, such as in the amygdala or brainstem^27^. On the other hand, there is a reported case of LRRK2-associated parkinsonism that did not demonstrate postmortem Lewy pathology but who demonstrated asyn aggregates on brain homogenate by asyn SAA^28^. Regardless of detection of asyn, of course, this does not exclude the possibility that pathogenic variants in LRRK2 may impact asyn function without leading to Lewy pathology or abnormal CSF SAA^29^.

It is also likely that in some individuals neurodegeneration does occur independent of presence of misfolded asyn. Indeed, pathogenic LRRK2 variants have been associated with various proteinopathies including AD, various tauopathies including PSP, corticobasal degeneration (CBD), familial frontotemporal degeneration (FTD), TDP-43-associated neurodegeneration^29^. Neuropathological studies are skewed toward cases with clinical features of parkinsonism^3^, and given that the prevalence of LRRK2 pathogenic variants in the general population is not small, interpretation of results in cases with other clinical diagnoses who have been autopsied is difficult; in some cases the genetic variant may be incidental. Having said that, a few studies that have screened for LRRK2 pathogenic variants in brain banks offer insights into the prevalence of LRRK2 pathogenic variants in a range of neurodegenerative disorders. In a series of 110 cases^30^, of which 66 were synucleinopathies, 29 tauopathies, and 3 non-specific nigral degeneration, the prevalence of positivity of pathogenic variants in LRRK2 gene was 1.8%. One case had PD based on clinical criteria and neuropathological examination, whereas another case had been diagnosed with PD based on clinical criteria, but neuropathological examination demonstrated nonspecific nigral degeneration without Lewy bodies. A p.R1441R variant was detected in another PD case^30^. Taking together data from published case series, approximately 22% of LRRK2 associated parkinsonism cases demonstrate neuropathological findings of hyperphosphorylated tau, as occurs in PSP^7^.

Several possible biologic mechanisms could be implicated in LRRK2-mediated neurodegeneration, whether related to asyn aggregates or independent of it. All pathogenic variants in the LRRK2 gene are missense mutations and have been found throughout the gene^29^.The LRRK2 protein is a large, complex multidomain protein that functions as a protein kinase. Altered LRRK2 signaling has been implicated in dysfunction in a range of cellular processes and molecular pathways including vesicular trafficking, autophagy, lysosomal degradation, endolysosomal stress, microglial response, calcium dysmetabolism and resultant endoplasmic reticulum stress, neuroinflammation, mitophagy, and mitochondrial dysfunction^5–7^. The PPMI cohort is being characterized with extensive proteomics and transcriptomics data which will allow investigation of differences in these various biologic processes in asyn positive and negative cases in the future.

### Female predominance among LRRK2 parkinsonism cases without evidence for asyn aggregates

We found a female predominance among the LRRK2 parkinsonism cases without evidence for pathologic asyn. There is extensive literature that demonstrates that in individuals diagnosed with PD, sex differences exist for clinical, biomarker, neuropathological, or genetic endpoints^31^. Sex differences in LRRK2-parkinsonism cases are particularly notable. A meta-analysis^32^ of 66 studies of LRRK2-associated parkinsonism (that were not biologically characterized) revealed a higher prevalence of LRRK2 pathogenic variants in females diagnosed with PD. In a study^33^ of 530 LRRK2-associated parkinsonism, and compared to 759 sporadic PD cases, the male predominance observed in sporadic PD was not seen in the LRRK2-associated cases.

As mentioned, asyn-negative LRRK2 parkinsonism cases often exhibit AD pathology, and these results could in part be a reflection of sex differences in AD. For example, women have a greater burden of neurofibrillary tangles^34,35^, and women with AD pathology are more likely to manifest clinically with dementia but not to be diagnosed with DLB^35^. The effect of sex on tau may even be brain-region specific, and females may have network characteristics favoring spread of tau^36^. Women with AD pathology are more likely to have copathology with TDP/hippocampal sclerosis abd cerebrovascular disease. On the other hand, male sex is more likely to be associated with pure Lewy body pathology (absence of copathology^34^).

There are several possible mechanisms that could explain sex differences in asyn pathology and in relation to LRRK2 that require investigation. Exposure to sex hormones has been postulated as one possible mechanism explaining a predominance of tau pathology in females compared to males. The higher likelihood of diagnosis of neurodegenerative disorders post-menopause has been observed. Estradiol may have a protective effect against hyperphosphorylation of tau^38^. Estrogen receptor colocalizes with neurofibrillary tangles.

Another possible mechanism may relate to the effect of estrogen on mitochondria and oxidative stress^38,39^. A relationship between sex hormones and neuroinflammation is another possible mechanism that may explain sex differences^39^, which is of particular relevance given the role of LRRK2 in the immune system. Sex differences in immune activation and microglial function may also play a role^31,39^.. Indeed, a study examining the serum profile of 23 immune-associated markers in sporadic and LRRK2-associated PD demonstrated sex differences in immune profile but without differences in the LRRK2 and sporadic group^40^. While differential genetic risk factors for PD in men vs women have not been demonstrated, sex-specific effects of genotype may exist^31,41^. 11 genomic loci have jointly been associated with PD and sex-specific traits, namely age of menarche and age at menopause. Many of the genes that mapped to loci shared between PD and age at menarche have been implicated in PD pathophysiology, including immune activation and regulation^41^. Sex-specific differences in LRRK2 brain expression in healthy controls (but not in PD) have been observed^41^. The effect of age on expression of genes that may be relevant in PD pathophysiology may also vary by sex^41^.. A greater burden of tau among women has been postulated to be mediated by ApoE status^42^, and upregulation of ApoE expression by estrogen was postulated as a possible mechanism^42^.

We did not find differences in ApoEe4 genotype in CSFasynSAA+ vs CSFasynSAA-but we had a small sample size and low prevalence of ApoEe4 in our sample. With larger sample sizes and by comparing proteomic or transcriptomic data, these hypotheses can be investigated in future studies. Gender differences in behavioral, occupational, environmental exposures may also contribute^7,43,44^ and deserve investigation.

### Lower Prevalence of Olfactory Dysfunction in the group without evidence for asyn aggregates, especially among females

A lower prevalence of olfactory deficit among LRRK2-associated parkinsonism has been previously identified^33,45^, but in PPMI it has been demonstrated that this finding is largely restricted to LRRK2-associated parkinsonism without evidence of asyn pathology^4^. In a study^33^ of 530 LRRK2-associated parkinsonism, and compared to 759 sporadic PD cases, female LRRK2-parkinsonism individuals were less likely to have olfactory deficit^33^.

However, in that study, biological characterization was not present. The PPMI study sample now enables demonstration that asyn negative LRRK2 parkinsonism cases are much more likely to be normosmic^4^. One possible explanation for these findings is the preferential susceptibility of olfactory bulb^46^ and anterior olfactory nucleus^47,48^ cells to asyn pathology, as evidenced by data from animal models. Future studies of asyn pathology in nasal mucosa in LRRK2 cases may shed light on the observed differences in olfactory dysfunction we report here.

### Less Severe Motor Dysfunction and Functional Impairment in the group without evidence for asyn aggregates

Despite being older, having similar disease duration, and lower LEDD at baseline assessment, the CSFasynSAA-had similar scores on MDS-UPDRS including part III ON score to the CSFasynSAA+ group. These results may indicate less severe motor involvement in the CSFasynSAA-group. Concordant with this, the CSFasynSAA-group remained stable in the MDS-UPDRS II, a multidomain, motor-predominant functional rating scale, whereas the CSFasynSAA+_group had a significant increase (declining function) over time. One possibility to explain these findings is that the underlying pathology in these cases leads to less severe affectation of dopaminergic pathways and other pathways implicated in parkinsonian motor abnormalities. The less severe DAT loss in this group supports this hypothesis. Indeed, while dopaminergic neuronal loss occurs in a range of neurodegenerative disorders, there may be disease-specific susceptibility.

In light of the differences in MDS-UPDRS II in the LRRK2-parkinsonim CSFasynSAA-vs CSFasynSAA+ cases, we next examined differences in LRRK2 CSFasynSAA-vs sPD CSFasynSAA+, as this analysis can provide insights as to whether the differences are unique to LRRK2 parkinsonism or are rather more a reflection of asyn aggregates status. Some differences in CSFasynSAA– and CSFasynSAA+ cases persisted, indicating that the differences may not be unique to LRRK2, though LRRK2 may still mediate some of these differences.

Prior studies have suggested that individuals with LRRK2-parkinsonism may be less likely to demonstrate motor complications compared to sporadic PD cases, especially among females^33^. However, in those studies biologic characterization was not available^33^. Our findings indicate that the more benign phenotype in LRRK2-associated parkinsonism may be driven by asyn-negative cases, A comparison of LRRK2-parkinsonism cases with asyn aggregates to sporadic cases with asyn aggregates is needed to determine the influence of the pathogenic variant itself on phenotype among those with asyn aggregates, and this analysis is underway in the PPMI cohort.

### Differences in non-motor features in those with vs without evidence for asyn aggregates

In the few available studies that compared clinical features in LRRK2-associated cases according to asyn status, a few clinical differences have been described^2^. Kalia et al demonstrated that among cases of LRRK2-associated parkinsonism, some non-motor symptoms associated with typical sporadic PD such as anxiety, orthostasis, and cognitive changes are more likely in those with evidence of asyn aggregates^2^.

In contrast, we found that the CSFasynSAA-group had greater global cognitive dysfunction, as assessed with MoCA, at baseline. These results should be interpreted with caution given the age, sex, and education differences in the two groups; indeed, results were no longer significant after adjusting for these possible confounders. Similarly, MoCA score was lower in the CSFasynSAA-LRRK2 group compared to the CSFasynSAA+ sPD group, but given the differences in age, sex, education, and disease duration despite frequency-matching, the significance of these results is unclear. Nevertheless, it remains possible that CSFasynSAA-LRRK2 parkinsonism cases are at risk for greater cognitive dysfunction. Given that such cases may be more likely to have tauopathy-mediated neurodegeneration, and it is possible that tau-based neurodegeneration affects cortical structures preferentially leading to greater cognitive impairment. Possibly supporting this hypothesis is the finding that total tau and phospho tau levels were higher in the CSFasynSAA-group, though this finding did not remain significant when adjusting for age. Further, the CSFasynSAA– and CSFasynSAA+ groups progressed similarly in terms of cognitive decline.

There was some indication that the rate of change in autonomic symptoms differed in the CSFasynSAA+ and CSFasynSAA-groups; differences in dysautonomia according to presence of asyn has also been reported by Kalia et al^2^. However, the clinical relevance of the findings in our study is not clear; the overall burden of autonomic symptoms was similar in the two groups and mean group scores did not change substantially over time.

### Biomarker differences: higher DAT binding and higher serum nFL

When examining DAT binding quantitatively, the CSFasynSAA-group had higher putamen DAT binding compared to the CSFasynSAA+ group. While the explanation for this is unclear, it may suggest that the neurodegenerative processes in CSFasynSAA-vs CSFasynSAA+ cases differentially affect dopaminergic neurons.

The CSFasynSAA-group had higher serum NfL. Serum NfL is a nonspecific marker of neuro-axonal injury and degeneration that may be abnormal in a range of neurologic disorders including FTD, multiple system atrophy (MSA), amyotrophic lateral sclerosis, stroke, multiple sclerosis, and traumatic brain injury among others^49^. It is higher in individuals diagnosed with the atypical parkinsonian disorders such as MSA and PSP compared to PD^50^. Across diseases, including in individuals diagnosed with PD, DLB, and AD, higher serum NfL is associated with greater cognitive dysfunction and predicts cognitive decline^19,51^. Consistent with this, in our study, the CSFasynSAA-group had lower MoCA at baseline, even after adjusting for age. However, the CSF asyn CSFasynSAA-group did not progress more on cognitive measures over time compared to the CSFasynSAA+ group. It is possible our study was underpowered to detect differences in longitudinal change over just a 4-year follow-up period. Alternatively, distinct biological mechanisms may subserve the progression on cognitive function.

### Genotype-phenotype correlations

While the majority of our sample carried the p.G2019S pathogenic variant, 14% had other variants, and there was a predominance of p.R1441G in the CSFasynSAA-group. These results are consistent with findings from the literature, mainly from neuropathologically examined case series^1–3,5,29,52^. Among 42 G2019S cases and 27 cases with other LRRK2 variants, the majority of G2019S carriers, 70-80%, have Lewy bodies, whereas only 40-45% of other LRRK2 variants do^1^. In the original family in which the LRRK2 locus was identified as being associated with parkinsonism^53^, and in the few subsequently examined cases now known to the I2020T variant, the pathology demonstrated pure nigral degeneration in the absence of Lewy bodies or neurofibrillary tangles in about 50% of cases. Tau pathology also varies according to genotype; 90% of neuropathologically examined G2019S LRRK2-parkinsonism cases have tau pathology compared to 38% of cases with other variants^5^. Importantly, among individuals carrying the same variant, even within a family, clinical and neuropathologic phenotypic variation exists^54^. For example, in a kindred of 4 cases with R1441C mutation, parkinsonism and nigral cell loss with depigmentation and gliosis of the substantia nigra pars compacta, 2 had Lewy bodies and 1 did not have asyn pathology but had neurofibrillary tangles. One hypothesis is that the specific genetic changes alter LRRK2 protein function differently. The p.G2019S and p.I2020T variants are in the kinase domain, and are known to increase LRRK2 kinase activity^5^. On the other hand, the p.R1441G/C/H/S pathogenic variant are in the ROC domain, and increase LRRK2 kinase activity by affecting GTPase function^5^. Pathogenic variants in parts of the gene that encode any of the 3 core catalytic domains of the LRRK2 protein, namely the Roc, COR, or kinase, can be associated with nigral degeneration without asyn pathology. However, available data indicate that p.R1441C/G/H, p.Y1699C and PI2020T are more likely than G2019S to be asyn negative^2,7,55^. In rare cases, pathological findings are consistent with MSA^7^.

### Genetic modifiers: comparison of PD risk variants in those with vs without evidence of asyn aggregates

To identify possible genetic underpinnings associated with CSF asyn SAA status, we compared PD risk variants in the groups. Previous research has estimated the heritability of PD at 22%, with PRS explaining approximately a quarter of this heritability within the European population^16^. Furthermore, PRS has been linked to an elevated risk of PD in carriers of the LRRK2 p.G2019S mutation, particularly noting a stronger association in cases of early-onset LRRK2-parkinsonism^44^. Interestingly, variants in MAPT^5^ have been reported to increase risk of PD in LRRK2 variant carriers. Other genetic variants that may modify manifestations of LRRK2 are in the DNM3 and VAMP4 genes^5,56^. Our investigation aimed to ascertain whether a correlation exists between mPRS and CSF asyn SAA status among LRRK2 parkinsonism cases. We did not find differences in mPRS between the groups, nor in the aforementioned genes. However, in analysis of individual risk variants, 3 were identified as possibly associated with CSF asyn SAA status, rs11557080, rs12951632, and rs6808178. Although they would not withstand correction for multiple testing, the variant rs11557080, located in the 3’ UTR of RAB29, is of particular interest due to previous studies suggesting an interaction between RAB29 and LRRK2 activity^57^.

Many studies that have investigated genetic modifiers in LRRK2-associated parkinsonism did not account for underlying pathology. In future studies, stratification of manifest cases according to evidence of asyn aggregates may yield new insights.

## Study Limitations

Strengths of this study include the large sample size, in vivo assessment of asyn with a robustly validated assay, and extensive clinical and biomarker characterization of the sample longitudinally. We limited our analysis to 4-years follow-up and are not able to draw conclusions on longer-term differences in the two groups. The biomarkers compared in this analysis reflect currently available analytes in the PPMI study. They provide limited insight into potential pathogenic mechanisms that may or may not diverge in LRRK2 parkinsonism CSFasynSAA-vs SAA+ cases. However, PPMI has a comprehensive biofluids repository that will further allow exploration of other biomarkers as they are validated. Due to the small sample size of participants with non-G2019S variants, we cannot draw conclusions regarding genotype-phenotype differences.

## Conclusion

In this study, we have demonstrated several characteristics that are different in LRRK2 associated parkinsonism cases with vs without evidence of asyn aggregates in the CSF. LRRK2-associated parkinsonism cases without asyn aggregates are more likely to be female, normosmic, to have relatively milder motor manifestations, and to exhibit less functional decline. They may also exhibit greater cognitive impairment. We demonstrate important biomarker differences including less loss of DAT binding and higher serum NfL in the CSF asyn negative group. The PPMI cohort is being characterized with extensive proteomics and transcriptomics data. This will allow investigation of differences in various biologic processes in LRRK2-associated parkinsonism asyn positive and negative cases in the future.

## Data availability

Data used in the preparation of this article were obtained on January 8, 2024 from the Parkinson’s Progression Markers Initiative (PPMI) database (www.ppmi-info.org/access-data-specimens/download-data), RRID:SCR 006431. For up-to-date information on the study, visit www.ppmi-ifo.org.

Statistical analysis codes used to perform the analyses in this article are shared on Zenodo https://doi.org/10.5281/zenodo.12682377.

Data Tier: This analysis was conducted by the PPMI Statistics Core and used actual dates of activity for participants, a restricted data element not available to public users of PPMI data.

## Funding

PPMI – a public-private partnership – is funded by the Michael J. Fox Foundation for Parkinson’s Research and funding partners, including 4D Pharma, Abbvie, AcureX, Allergan, Amathus Therapeutics, Aligning Science Across Parkinson’s, AskBio, Avid Radiopharmaceuticals, BIAL, BioArctic, Biogen, Biohaven, BioLegend, BlueRock Therapeutics, Bristol-Myers Squibb, Calico Labs, Capsida Biotherapeutics, Celgene, Cerevel Therapeutics, Coave Therapeutics, DaCapo Brainscience, Denali, Edmond J. Safra Foundation, Eli Lilly, Gain Therapeutics, GE HealthCare, Genentech, GSK, Golub Capital, Handl Therapeutics, Insitro, Jazz Pharmaceuticals, Johnson & Johnson Innovative Medicine, Lundbeck, Merck, Meso Scale Discovery, Mission Therapeutics, Neurocrine Biosciences, Neuron23, Neuropore, Pfizer, Piramal, Prevail Therapeutics, Roche, Sanofi, Servier, Sun Pharma Advanced Research Company, Takeda, Teva, UCB, Vanqua Bio, Verily, Voyager Therapeutics, the Weston Family Foundation and Yumanity Therapeutics.

This work was supported in part by the Intramural Research Program of the National Institute on Aging (NIA) and the Center for Alzheimer’s and Related Dementias (CARD) under Award Number AG000534

H.I.’s participation in this project was part of a competitive contract awarded to DataTecnica LLC by the National Institutes of Health to support open science research.

## Competing interests

The authors report no competing interests.

## Supplementary material

Supplementary Table 1, Supplementary Table 2

## Abbreviations

AD: Alzheimer’s Disease
AMP-PD: Accelerating Medicines Partnership Parkinson’s Disease
Asyn: Alpha-synuclein
BMP: Bis(monoacylglycero)phosphate
CBD: Corticobasal Degeneration
CSF: Cerebrospinal fluid
CSFasynSAA+: Positive CSF asyn SAA
CSFasynSAA−: Negative CSF asyn SAA
DAT: Dopamine transporter binding
DLB: Dementia with Lewy Bodies
ESS: Epworth Sleepiness Scale
FTD: Familial Frontotemporal Degeneration
GDS-15: Geriatric Depression Scale-15 item
GWAS: Genome-wide Association Studies
HVLT-R: Hopkins Verbal Learning Test-Revised
LEDD: Levodopa equivalent daily dose
LMM: Linear mixed-effects models
MAR: Missing at random
MCI: Mild Cognitive Impairment
MDS-UPDRS: Movement Disorders Society Modified Unified Parkinson’s Disease Rating Scale
MoCA: Montreal Cognitive Assessment
mPRS: Modified polygenic risk score
MSA: Multiple System Atrophy
Nfl: Neurofilament light
PD: Parkinson’s Disease
PPMI: Parkinson Progression Markers Initiative
PRS: Polygenic risk score
PSP: Progressive Supranuclear Palsy
QUIP-RS: Questionnaire for Impulsive-Compulsive Disorders in Parkinson’s Disease–Rating Scale
RBD: Rapid Eye Movement Behavior Disorder
SAA: Seed amplification assay
SBR: Specific binding ratio
SCOPA-AUT: Scales for Outcomes in Parkinson’s-Autonomic
SNP: Single-nucleotide polymorphism
sPD: Sporadic Parkinson’s Disease
SPECT: Single-photon emission computed tomography
STAI: State and Trait Anxiety Scale
TDP-43: TAR DNA-binding protein 43
UPSIT: University of Pennsylvania Smell Identification Test

## Appendix 1

### Executive Steering Committee

Kenneth Marek, MD^1^ (Principal Investigator); Caroline Tanner, MD, PhD^9^; Tanya Simuni, MD^3^; Andrew Siderowf, MD, MSCE^12^; Douglas Galasko, MD^27^; Lana Chahine, MD^39^; Christopher Coffey, PhD^4^; Kalpana Merchant, PhD^59^; Kathleen Poston, MD^38^; Roseanne Dobkin, PhD^41^; Tatiana Foroud, PhD^15^; Brit Mollenhauer, MD^8^; Dan Weintraub, MD^12^; Ethan Brown, MD^9^; Karl Kieburtz, MD, MPH^23^; Mark Frasier, PhD^6^; Todd Sherer, PhD^6^; Sohini Chowdhury, MA^6^; Roy Alcalay, MD^35^ and Aleksandar Videnovic, MD^45^

### Steering Committee

Duygu Tosun-Turgut, PhD^9^; Werner Poewe, MD^7^; Susan Bressman, MD^14^; Jan Hammer^15^; Raymond James, RN^22^; Ekemini Riley, PhD^40^; John Seibyl, MD^1^; Leslie Shaw, PhD^12^; David Standaert, MD, PhD^18^; Sneha Mantri, MD, MS^60^; Nabila Dahodwala, MD^12^; Michael Schwarzschild^45^; Connie Marras^43^; Hubert Fernandez, MD^25^; Ira Shoulson, MD^23^; Helen Rowbotham^2^; Paola Casalin^11^ and Claudia Trenkwalder, MD^8^

### Michael J. Fox Foundation (Sponsor)

Todd Sherer, PhD; Sohini Chowdhury, MA; Mark Frasier, PhD; Jamie Eberling, PhD; Katie Kopil, PhD; Alyssa O’Grady; Maggie McGuire Kuhl; Leslie Kirsch, EdD and Tawny Willson, MBS

### Study Cores, Committees and Related Studies

*Project Management Core:* Emily Flagg, BA^1^

*Site Management Core:* Tanya Simuni, MD^3^; Bridget McMahon, BS^1^

Strategy and Technical Operations: Craig Stanley, PhD^1^; Kim Fabrizio, BA^1^

*Data Management Core:* Dixie Ecklund, MBA, MSN^4^; Trevis Huff, BSE^4^

*Screening Core:* Tatiana Foroud, PhD^15^; Laura Heathers, BA^15^; Christopher Hobbick, BSCE^15^; Gena Antonopoulos, BSN^15^

*Imaging Core:* John Seibyl, MD^1^; Kathleen Poston, MD^38^

*Statistics Core*: Christopher Coffey, PhD^4^; Chelsea Caspell-Garcia, MS^4^; Michael Brumm, MS^4^

*Bioinformatics Core*: Arthur Toga, PhD^10^; Karen Crawford, MLIS^10^

*Biorepository Core:* Tatiana Foroud, PhD^15^; Jan Hamer, BS^15^

*Biologics Review Committee*: Brit Mollenhauer^8^; Doug Galasko^27^; Kalpana Merchant^59^

*Genetics Core:* Andrew Singleton, PhD^13^

*Pathology Core:* Tatiana Foroud, PhD^15^; Thomas Montine, MD, PhD^38^

*Found:* Caroline Tanner, MD PhD^9^

*PPMI Online:* Carlie Tanner, MD PhD^9^; Ethan Brown, MD^9^; Lana Chahine, MD^39^; Roseann Dobkin, PhD^41^; Monica Korell, MPH^9^

### Site Investigators

Charles Adler, PhD^49^; Roy Alcalay, MD^35^; Amy Amara, PhD^50^; Paolo Barone, PhD^30^; Bastiaan Bloem, PhD^58^ Susan Bressman, MD^14^; Kathrin Brockmann, MD^26^; Norbert Brüggemann, MD^57^; Lana Chahine, MD^39^; Kelvin Chou, MD^42^; Nabila Dahodwala, MD^12^; Alberto Espay, MD^32^; Stewart Factor, DO^16^; Hubert Fernandez, MD^25^; Michelle Fullard, MD^50^; Douglas Galasko, MD^27^; Robert Hauser, MD^19^; Penelope Hogarth, MD^17^; Shu-Ching Hu, PhD^21^; Michele Hu, PhD^56^; Stuart Isaacson, MD^31^; Christine Klein, MD^57^; Rejko Krueger, MD^2^; Mark Lew, MD^47^; Zoltan Mari, MD^54^; Connie Marras, PhD^43^; Maria Jose Martí, PhD^33^; Nikolaus McFarland, PhD^52^; Tiago Mestre, PhD^44^; Brit Mollenhauer, MD^8^; Emile Moukheiber, MD^28^; Alastair Noyce, PhD^61^ Wolfgang Oertel, PhD^62^; Njideka Okubadejo, MD^63^; Sarah O’Shea, MD^37^; Rajesh Pahwa, MD^46^; Nicola Pavese, PhD^55^; Werner Poewe, MD^7^; Ron Postuma, MD^53^; Giulietta Riboldi, MD^51^; Lauren Ruffrage, MS^18^; Javier Ruiz Martinez, PhD^34^; David Russell, PhD^1^; Marie H Saint-Hilaire, MD^22^; Neil Santos, BS^49^; Wesley Schlett^45^; Ruth Schneider, MD^23^; Holly Shill, MD^48^; David Shprecher, DO^24^; Tanya Simuni, MD^3^; David Standaert, PhD^18^; Leonidas Stefanis, PhD^36^; Yen Tai, PhD^29^; Caroline Tanner, PhD^9^; Arjun Tarakad, MD^20^; Eduardo Tolosa PhD^33^ and Aleksandar Videnovic, MD^45^

### Coordinators

Susan Ainscough, BA^30^; Courtney Blair, MA^18^; Erica Botting^19^; Isabella Chung, BS^54^; Kelly Clark^24^; Ioana Croitoru^34^; Kelly DeLano, MS^32^; Iris Egner, PhD^7^; Fahrial Esha, BS^51^; May Eshel, MSc^35^; Frank Ferrari, BS^42^; Victoria Kate Foster^55^; Alicia Garrido, MD^33^; Madita Grümmer^57^; Bethzaida Herrera^48^; Ella Hilt^26^; Chloe Huntzinger, BA^50^; Raymond James, BS^22^; Farah Kausar, PhD^9^; Christos Koros, MD, PhD^36^; Yara Krasowski, MSc^58^; Dustin Le, BS^17^; Ying Liu, MD^50^; Taina M. Marques, PhD^2^; Helen Mejia Santana, MA^37^; Sherri Mosovsky, MPH^39^; Jennifer Mule, BS^25^; Philip Ng, BS^43^; Lauren O’Brien^46^; Abiola Ogunleye, PGDip^29^; Oluwadamilola Ojo, MD^63^; Obi Onyinanya, BS^28^; Lisbeth Pennente, BA^31^; Romina Perrotti^53^; Michael Pileggi, MS^53^; Ashwini Ramachandran, MSc^12^; Deborah Raymond, MS^14^; Jamil Razzaque, MS^56^; Shawna Reddie, BA^44^; Kori Ribb, BSN,^28^; Kyle Rizer, BA^52^; Janelle Rodriguez, BS^27^; Stephanie Roman, HS^1^; Clarissa Sanchez, MPH^20^; Cristina Simonet, PhD^29^; Anisha Singh, BS^23^; Elisabeth Sittig, RN^62^; Barbara Sommerfeld MSN^16^; Angela Stovall, BS^42^; Bobbie Stubbeman, BS^32^; Alejandra Valenzuela, BS^47^; Catherine Wandell, BS^21^; Diana Willeke^8^; Karen Williams, BA^3^ and Dilinuer Wubuli, MB^43^

1 Institute for Neurodegenerative Disorders, New Haven, CT
2 University of Luxembourg, Luxembourg
3 Northwestern University, Chicago, IL
4 University of Iowa, Iowa City, IA
5 VectivBio AG
6 The Michael J. Fox Foundation for Parkinson’s Research, New York, NY
7 Innsbruck Medical University, Innsbruck, Austria
8 Paracelsus-Elena Klinik, Kassel, Germany
9 University of California, San Francisco, CA
10 Laboratory of Neuroimaging (LONI), University of Southern California
11 BioRep, Milan, Italy
12 University of Pennsylvania, Philadelphia, PA
13 National Institute on Aging, NIH, Bethesda, MD
14 Mount Sinai Beth Israel, New York, NY
15 Indiana University, Indianapolis, IN
16 Emory University of Medicine, Atlanta, GA
17 Oregon Health and Science University, Portland, OR
18 University of Alabama at Birmingham, Birmingham, AL
19 University of South Florida, Tampa, FL
20 Baylor College of Medicine, Houston, TX
21 University of Washington, Seattle, WA
22 Boston University, Boston, MA
23 University of Rochester, Rochester, NY
24 Banner Research Institute, Sun City, AZ
25 Cleveland Clinic, Cleveland, OH
26 University of Tübingen, Tübingen, Germany
27 University of California, San Diego, CA
28 Johns Hopkins University, Baltimore, MD
29 Imperial College of London, London, UK
30 University of Salerno, Salerno, Italy
31 Parkinson’s Disease and Movement Disorders Center, Boca Raton, FL
32 University of Cincinnati, Cincinnati, OH
33 Hospital Clinic of Barcelona, Barcelona, Spain
34 Hospital Universitario Donostia, San Sebastian, Spain
35 Tel Aviv Sourasky Medical Center, Tel Aviv, Israel
36 National and Kapodistrian University of Athens, Athens, Greece
37 Columbia University Irving Medical Center, New York, NY
38 Stanford University, Stanford, CA
39 University of Pittsburgh, Pittsburgh, PA
40 Center for Strategy Philanthropy at Milken Institute, Washington D.C.
41 Rutgers University, Robert Wood Johnson Medical School, New Brunswick, New Jersey
42 University of Michigan, Ann Arbor, MI
43 Toronto Western Hospital, Toronto, Canada
44 The Ottawa Hospital, Ottawa, Canada
45 Massachusetts General Hospital, Boston, MA
46 University of Kansas Medical Center, Kansas City, KS
47 University of Southern California, Los Angeles, CA
48 Barrow Neurological Institute, Phoenix, AZ
49 Mayo Clinic Arizona, Scottsdale, AZ
50 University of Colorado, Aurora, CO
51 NYU Langone Medical Center, New York, NY
52 University of Florida, Gainesville, FL
53 Montreal Neurological Institute and Hospital/McGill, Montreal, QC, Canada
54 Cleveland Clinic-Las Vegas Lou Ruvo Center for Brain Health, Las Vegas, NV
55 Clinical Ageing Research Unit, Newcastle, UK
56 John Radcliffe Hospital Oxford and Oxford University, Oxford, UK
57 Universität Lübeck, Luebeck, Germany
58 Radboud University, Nijmegen, Netherlands
59 TransThera Consulting
60 Duke University, Durham, NC
61 Wolfson Institute of Population Health, Queen Mary University of London, UK
62 Philipps-University Marburg, Germany
63 University of Lagos, Nigeria

## Notes

### Competing Interest Statement

The authors have declared no competing interest.

### Clinical Protocols

https://dx.doi.org/10.17504/protocols.io.n92ldmw6ol5b/v2

