## Supplement Tables for "LRRK2-Associated Parkinsonism With and Without *In Vivo* Evidence of Alpha-Synuclein Aggregates"

***Supplementary Table 1. Results of linear mixed effects models***

|  |  | ***Linear Assumption*** | | | ***Quadratic Assumption*** | | ***3-way Sex, Time, & SAA Interaction*** | |
| --- | --- | --- | --- | --- | --- | --- | --- | --- |
|  | ***Variable*** | ***Interaction***  ***p-value*** | ***Time effect estimate*** | ***Time effect p-value*** | ***p-value*** | ***Effect*** | ***p-value*** | ***Effect*** |
| 1 | **mS&E** | 0.894 | -0.736 (-1.502, 0.029) | 0.059 | 0.097 | -1.309 | 0.135 | -2.371 |
| 2 | **HY stage (>2) - ON** | 0.120 | 1.376 (0.846, 2.239) | 0.198 | 0.573 | -0.342 | 0.149 | 2.112 |
| 3 | **MDS-UPDRS I** | 0.637 | 0.436 (0.119, 0.752) | 0.007 | 0.133 | 0.449 | 0.265 | 0.728 |
| 4 | **MDS-UPDRS II** | 0.037 | POS: 0.837 (0.467, 1.207) NEG: 0.108 (-0.466, 0.682) | Group POS: <.001 Group NEG: 0.711 | 0.003 | 1.109 | 0.199 | 0.915 |
| 5 | **MDS-UPDRS III - ON** | 0.960 | 0.504 (-0.442, 1.450) | 0.294 | 0.923 | -0.075 | 0.396 | -1.649 |
| 6 | **Total MDS-UPDRS - ON** | 0.722 | 1.140 (-0.246, 2.527) | 0.106 | 0.115 | 1.743 | 0.744 | -0.932 |
| 7 | **Ambulatory Capacity Score - ON** | 0.262 | 0.064 (-0.094, 0.223) | 0.424 | 0.227 | 0.206 | 0.607 | 0.166 |
| 8 | **Geriatric Depression Scale** | 0.182 | 0.104 (-0.091, 0.299) | 0.295 | 0.946 | -0.012 | 0.843 | -0.081 |
| 9 | **State-Trait Anxiety Inventory** | 0.589 | 0.617 (-0.603, 1.837) | 0.319 | 0.846 | -0.201 | 0.488 | 1.762 |
| 10 | **SCOPA-AUT** | 0.927 | 0.468 (-0.121, 1.057) | 0.118 | 0.003 | -1.317 | 0.333 | 1.185 |
| 11 | **REM Sleep Behavior Disorder** | 0.565 | 0.067 (-0.086, 0.220) | 0.387 | 0.352 | 0.149 | 0.518 | 0.207 |
| 12 | **REM Sleep Behavior Disorder (>6)** | 0.863 | 1.256 (0.901, 1.751) | 0.179 | 0.762 | 0.115 | 0.885 | 0.107 |
| 13 | **Epworth Sleepiness Scale** | 0.524 | 0.041 (-0.198, 0.280) | 0.736 | 0.825 | 0.051 | 0.394 | 0.425 |
| 14 | **Montreal Cognitive Assessment** | 0.762 | 0.110 (-0.065, 0.284) | 0.217 | 0.403 | -0.130 | 0.203 | 0.462 |
| 15 | **Benton Judgement of Line Orientation scaled score** | 0.559 | 0.079 (-0.132, 0.290) | 0.461 | 0.942 | 0.017 | 0.420 | 0.352 |
| 16 | **HVLT Immediate/Total Recall t-score** | 0.545 | -0.315 (-1.132, 0.501) | 0.446 | 0.070 | 1.406 | 0.442 | 1.304 |
| 17 | **Letter Number Sequencing Score scaled score** | 0.420 | 0.061 (-0.113, 0.236) | 0.487 | 0.513 | -0.112 | 0.906 | -0.043 |
| 18 | **Semantic Fluency Total Score t-score** | 0.191 | -0.295 (-0.993, 0.403) | 0.404 | 0.906 | 0.092 | 0.537 | 0.892 |
| 19 | **Number of ICDs** | 0.202 | 0.988 (0.770, 1.267) | 0.924 | 0.443 | -0.220 | 0.472 | -0.380 |
| 20 | **DAT SBR lowest putamen** | 0.919 | -0.011 (-0.020, -0.002) | 0.021 |  |  | 0.199 | 0.024 |
| 21 | **CSF abeta*** | 0.625 | 0.324 (-3.977, 4.624) | 0.882 | 0.814 | -1.044 | 0.160 | -12.391 |
| 22 | **CSF abeta <= 683** | 0.615 | 1.005 (0.604, 1.674) | 0.985 | 0.472 | -0.395 | 0.826 | 0.234 |
| 23 | **CSF abeta <= 710** | 0.580 | 1.016 (0.617, 1.675) | 0.949 | 0.444 | -0.412 | 0.755 | 0.327 |
| 24 | **CSF tau*** | 0.239 | 2.316 (-2.400, 7.031) | 0.333 | 0.438 | 3.787 | 0.610 | 4.885 |
| 25 | **CSF tau >= 266** | 0.695 | 0.916 (0.520, 1.614) | 0.761 | 0.901 | 0.075 | 0.114 | -2.038 |
| 26 | **CSF tau >=148** | 0.965 | 0.776 (0.493, 1.221) | 0.271 | 0.192 | 0.675 | 0.444 | 0.718 |
| 27 | **CSF ptau*** | 0.566 | 3.283 (0.202, 6.365) | 0.037 | 0.093 | 5.287 | 0.994 | -0.045 |
| 28 | **CSF ptau >= 24** | 0.478 | 0.688 (0.293, 1.616) | 0.388 |  |  | 0.162 | -3.191 |
| 29 | **CSF ptau >= 13** | 0.408 | 1.372 (0.899, 2.093) | 0.142 | 0.821 | -0.100 | 0.458 | 0.639 |
| 30 | **CSF tau-abeta Ratio*** | 0.255 | 3.534 (-2.591, 9.659) | 0.256 | 0.673 | 2.687 | 0.817 | -2.907 |
| 31 | **Serum NfL** | 0.613 | -3.491 (-29.168, 22.187) | 0.788 | 0.292 | -48.677 | 0.985 | -0.968 |
| 32 | **Total di-18:1 BMP** | 0.330 | 0.991 (-0.956, 2.939) | 0.313 |  |  | 0.152 | -5.692 |
| 33 | **Total di-22:6-BMP** | 0.307 | -4.358 (-12.773, 4.056) | 0.304 |  |  | 0.711 | 6.349 |
| 34 | **2.2 di-22:6 BMP** | 0.753 | -1.310 (-8.549, 5.930) | 0.719 |  |  | 0.231 | 17.641 |

mS&E=modified Schwab and England
ICD=impulse control disorder
SCOPA-AUT=Scales for Outcomes in Parkinson’s Disease - Autonomic Dysfunction
DAT SBR lowest putamen=dopamine transporter specific binding ratio, percent expected for age and sex, lowest of the right or left putamen values

*Scores were imputed with their upper and lower limits of detection. Results based on rank-based models. The interpretation of the estimates for rank-based models should be approached with caution, as they reflect changes in mean rank rather than changes in mean raw values.

***Supplementary Table 2: Longitudinal assessment of imaging and biofluid biomarkers***

| ***Variable*** |  | ***Baseline*** | ***Year 1*** | ***Year 2*** | ***Year 3*** | ***Year 4*** |
| --- | --- | --- | --- | --- | --- | --- |
| **DAT SBR lowest putamen** | | | | | | |
| N | SAA+ | 89 | 0 | 62 | 0 | 51 |
|  | SAA- | 45 | 0 | 31 | 0 | 24 |
| Mean (SD) | SAA+ | 0.29 (0.10) | N/A | 0.25 (0.10) | N/A | 0.22 (0.09) |
|  | SAA- | 0.38 (0.12) | N/A | 0.33 (0.15) | N/A | 0.31 (0.12) |
| **CSF abeta*** | | | | | | |
| N | SAA+ | 92 | 68 | 50 | 41 | 29 |
|  | SAA- | 41 | 32 | 26 | 19 | 13 |
| Mean (SD) | SAA+ | 837.8 (304.1) | 896.6 (316.5) | 863.1 (328.5) | 843.4 (327.4) | 805.5 (325.1) |
|  | SAA- | 947.6 (330.8) | 971.0 (304.0) | 927.4 (304.1) | 911.7 (290.8) | 847.6 (264.9) |
| **CSF abeta <= 683** | | | | | | |
| Yes | SAA+ | 30 (33%) | 19 (28%) | 20 (40%) | 17 (41%) | 12 (41%) |
|  | SAA- | 11 (27%) | 6 (19%) | 5 (19%) | 4 (21%) | 4 (31%) |
| **CSF abeta <= 710** | | | | | | |
| Yes | SAA+ | 36 (39%) | 19 (28%) | 22 (44%) | 17 (41%) | 13 (45%) |
|  | SAA- | 11 (27%) | 6 (19%) | 5 (19%) | 4 (21%) | 4 (31%) |
| **CSF tau*** | | | | | | |
| N | SAA+ | 93 | 68 | 50 | 42 | 29 |
|  | SAA- | 42 | 32 | 26 | 19 | 13 |
| Mean (SD) | SAA+ | 156.5 (56.5) | 166.1 (62.8) | 161.8 (71.0) | 155.5 (56.4) | 142.0 (44.5) |
|  | SAA- | 190.1 (61.8) | 192.7 (69.0) | 204.2 (84.4) | 174.4 (36.5) | 173.2 (49.1) |
| **CSF tau >= 266** | | | | | | |
| Yes | SAA+ | 5 (5%) | 6 (9%) | 4 (8%) | 2 (5%) | 1 (3%) |
|  | SAA- | 6 (14%) | 4 (13%) | 5 (19%) | 0 (0%) | 1 (8%) |
| **CSF tau >= 148** | | | | | | |
| Yes | SAA+ | 46 (49%) | 37 (54%) | 24 (48%) | 18 (43%) | 9 (31%) |
|  | SAA- | 32 (76%) | 25 (78%) | 22 (85%) | 15 (79%) | 8 (62%) |
| **CSF ptau*** | | | | | | |
| N | SAA+ | 93 | 68 | 50 | 42 | 29 |
|  | SAA- | 42 | 32 | 26 | 19 | 13 |
| Mean (SD) | SAA+ | 13.3 (5.0) | 13.9 (5.4) | 13.6 (5.4) | 12.9 (4.9) | 12.2 (4.1) |
|  | SAA- | 16.2 (5.2) | 16.7 (6.1) | 17.3 (7.6) | 14.9 (3.6) | 14.4 (4.2) |
| **CSF ptau >= 24** | | | | | | |
| Yes | SAA+ | 5 (5%) | 4 (6%) | 3 (6%) | 2 (5%) | 0 (0%) |
|  | SAA- | 4 (10%) | 4 (13%) | 4 (15%) | 0 (0%) | 0 (0%) |
| **CSF ptau >= 13** | | | | | | |
| Yes | SAA+ | 43 (46%) | 32 (47%) | 23 (46%) | 17 (40%) | 10 (34%) |
|  | SAA- | 30 (71%) | 22 (69%) | 18 (69%) | 12 (63%) | 8 (62%) |
| **CSF tau-abeta Ratio*** | | | | | | |
| N | SAA+ | 92 | 68 | 50 | 41 | 29 |
|  | SAA- | 41 | 32 | 26 | 19 | 13 |
| Mean (SD) | SAA+ | 0.206 (0.117) | 0.202 (0.112) | 0.207 (0.136) | 0.204 (0.126) | 0.209 (0.166) |
|  | SAA- | 0.215 (0.087) | 0.214 (0.098) | 0.242 (0.129) | 0.214 (0.103) | 0.221 (0.089) |
| **Serum NfL** | | | | | | |
| N | SAA+ | 82 | 73 | 56 | 49 | 0 |
|  | SAA- | 35 | 33 | 24 | 21 | 0 |
| Mean (SD) | SAA+ | 12.48 (8.07) | 14.52 (8.15) | 58.28 (327.46) | 14.57 (9.41) | N/A |
|  | SAA- | 18.73 (8.75) | 18.63 (10.96) | 18.90 (6.09) | 17.38 (5.79) | N/A |
| **Total di-18:1 BMP** | | | | | | |
| N | SAA+ | 91 | 71 | 52 | 0 | 0 |
|  | SAA- | 38 | 33 | 21 | 0 | 0 |
| Mean (SD) | SAA+ | 16 (14) | 16 (12) | 17 (13) | N/A | N/A |
|  | SAA- | 21 (26) | 19 (13) | 19 (13) | N/A | N/A |
| **Total di-22:6-BMP** | | | | | | |
| N | SAA+ | 91 | 71 | 52 | 0 | 0 |
|  | SAA- | 38 | 33 | 21 | 0 | 0 |
| Mean (SD) | SAA+ | 75 (49) | 76 (50) | 76 (43) | N/A | N/A |
|  | SAA- | 87 (55) | 86 (44) | 89 (50) | N/A | N/A |
| **2.2 di-22:6 BMP** | | | | | | |
| N | SAA+ | 91 | 71 | 52 | 0 | 0 |
|  | SAA- | 38 | 33 | 21 | 0 | 0 |
| Mean (SD) | SAA+ | 59 (41) | 59 (42) | 58 (37) | N/A | N/A |
|  | SAA- | 70 (47) | 67 (39) | 72 (44) | N/A | N/A |

DAT SBR lowest putamen=dopamine transporter specific binding ratio, percent expected for age and sex, lowest of the right or left putamen values

*Scores were imputed with their upper and lower limits of detection.
